# Spike protein antibodies mediate the apparent correlation between SARS-CoV-2 nucleocapsid antibodies and neutralization test results

**DOI:** 10.1101/2021.04.08.21255143

**Authors:** Thomas Perkmann, Thomas Koller, Nicole Perkmann-Nagele, Miriam Klausberger, Mark Duerkop, Barbara Holzer, Boris Hartmann, Patrick Mucher, Astrid Radakovics, Maria Ozsvar-Kozma, Oswald F Wagner, Christoph J Binder, Helmuth Haslacher

## Abstract

**Objectives:** SARS-CoV-2 infection induces the formation of different antibodies. However, not all of which might prevent the virus from entering the cell, although their concentrations correlate with the titers of viral neutralization tests (NTs). Antibodies against the viral nucleocapsid (NC), e.g., can be classified as such. We aimed to prove the hypothesis that the apparent correlation between NC-antibody levels and NT-titers is mediated by simultaneously occurring antibodies against viral spike-protein components.

**Methods:** We included 64 individuals with previous SARS-CoV-2 infection (>14d after symptom onset). SARS-CoV-2 antibodies against the NC (Roche total antibody ECLIA, Abbott IgG CMIA) and spike-protein (Technozym RBD ELISA, DiaSorin S1/S2 CLIA) were measured, and neutralization tests were performed. The effect of spike-protein antibodies on the correlation between NC-antibodies and NT-titers was evaluated by partial correlation and mediation analyses.

**Results:** Both tested assays assessing antibodies against the NC correlated significantly with NT titers: Abbott ρ=0.742, P<0.0001; Roche ρ=0.365, P<0.01. However, when controlling the rank correlations for the presence of RBD or S1/S2 antibodies, correlation coefficients dropped to ρ=0.318/ρ=0.329 (P<0.05/P<0.01), respectively for Abbott and vanished for Roche. As a result, only a maximum of 11% of NT titer variability could be explained by NC-antibody levels.

**Conclusions:** Our data suggest that the apparent correlation between NC antibodies and NT titers is strongly mediated by co-occurring RBD antibody concentrations. To avoid falsely implied causal relationships, all correlation analyses of non-spike-associated antibody assays and neutralization assays should include a partial correlation analysis to exclude a possible mediator effect of spike-associated antibodies.

## Introduction

Since the start of vaccinations against SARS-CoV-2, antibodies directed against the virus have been in the spotlight. Although the virus contains several immunogenic compounds that cause antibody production, not all of them act as neutralizing antibodies, i.e. antibodies which have the capacity to prevent the virus from entering host cells.

Specifically, antibodies against the viral spike protein, which also contains the receptor-binding domain (RBD), exhibit neutralizing properties (1). Therefore, current vaccination strategies aim to achieve immunity using spike protein components.

Additionally, some authors have shown that antibodies against the nucleocapsid protein (NC) correlate with neutralization activity as well. However, these structures are not present on the viral surface (2-4). Consequently, it has been hypothesized that these are mere spurious correlations mediated by the co-occurrence of antibodies to RBD (2, 5).

## Methods

To support this hypothesis, we measured levels of antibodies binding to the viral nucleocapsid antigen in 64 individuals with previous SARS-CoV-2 infection (either PCR-confirmed or symptomatic close contacts; sampling >14 days after symptom onset). We used various CE-labeled immunoassays (Roche total antibody ECLIA, Abbott IgG MCIA) and antibodies to spike protein targets (Technozym RBD IgG ELISA, DiaSorin S1/S2 IgG CLIA). The sera’s functional ability to neutralize SARS-CoV-2 viruses was tested using a classical TCID50 assay (NT) as previously described (6) with the modification that the heat-treated sera were diluted 1:4 in triplicates in serum-free HEPES-buffered DMEM medium. To quantify neutralization titer, the reciprocal of the highest serum dilution that protected more than 50% of the cells from the cytopathic effect was used and was calculated according to Reed and Muench (7).

Recruiting procedures, pre-analytical workflows and analytical methods were reported earlier (8-10). Samples were either left-over materials from the General Hospital Vienna or donated by convalescent donors giving written informed consent (EK 2011/404). This evaluation is part of a study which was reviewed and approved by the ethics committee of the Medical University of Vienna (EK 2257/2020).

As data did not meet the assumptions for parametric testing, all statistical analyses were performed on ranks. To test whether a correlation is disturbed by a mediator (e.g. RBD or S1/S2 antibody concentrations), partial rank correlation tests and mediation analyses, according to Hayes (11) were performed. The latter uses ordinary least squares regression, whereby confidence intervals were computed by bootstrapping with 5000 samples. In brief, the mediation analysis quantifies the total effect of NC antibody levels on NT titers (c), as well as the direct effect of NC antibody levels on NT titers (c’) after substracting the mediation effect of co-occuring RBD antibody concentrations.

## Results

Both tested assays assessing antibodies against the NC correlated significantly with NT titers: Abbott ρ=0.742, P=2.2×10^−12^; Roche ρ=0.365, P=0.003. However, if the rank correlations were controlled for RBD antibodies’ presence, correlation coefficients dropped to ρ=0.318 (P=0.011, Abbott), ρ=0.032 (P=0.806, Roche). The same holds if antibodies against an S1/S2 combination antigen were kept constant: ρ=0.329 (P=0.008, Abbott), ρ=-0.101 (P=0.430, Roche). When RBD or S1/S2 antibody concentrations were not considered, up to 55% of NT titers’ variability could be explained by NC antibody concentrations (Abbott). However, after introducing RBD or S1/S2 antibody concentrations as a mediator variable, only a maximum of 11% of NT titer variability could be explained by NC antibody levels (see Figure 1). This association was also evident in a mediation analysis on ranks. Both NC assays significantly predicted NT titers (Abbott: B=0.74, P<0.0001; Roche: B=0.36, P=0.002). After including Technozym RBD IgG concentrations as a mediator into the model, the effect of Abbott NC IgG levels on NT titers were partly mediated (indirect effect ab=0.46, 95%CI: 0.27 – 0.73) and the relationship between Roche NC total antibody levels and NT titers was fully mediated (ab=0.34, 95%CI: 0.17 – 0.53) by the RBD IgG concentrations (see Figure 1). In a last step, we aimed to assess whether the correlation between RBD antibody concentrations and NT titers could be attenuated to the same extent by keeping NC antibody concentrations constant. Although controlling for Abbott NC IgG-levels affected the coefficients of determination, the remaining R^2^ was still 0.31 for S1/S2 and 0.35 for RBD antibodies (see Figure 2).

**Figure 1:**
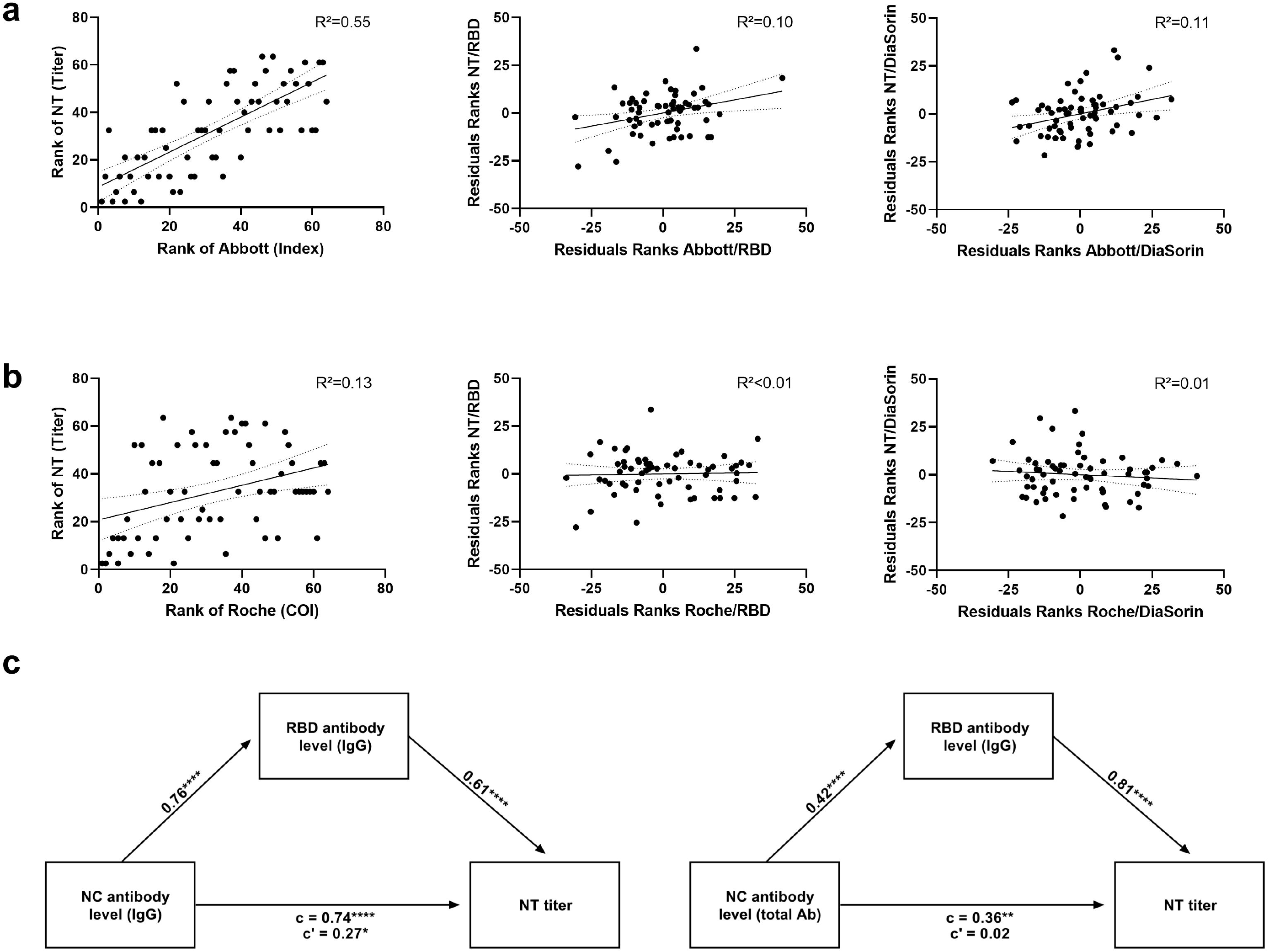
Rank correlations between titers of viral neutralization tests (NT) and nucleocapsid antibody concentrations measured by the a) Abbott CMIA (IgG) or the b) Roche ECLIA (IgG, IgM, IgA). The figures in the second column show the changes in rank correlations by keeping the RBD (Technozym) or S1/S2 (DiaSorin) IgG antibody concentration constant, respectively. c) mediation analysis to assess whether the effect of Abbott nucleocapsid (NC) IgG levels (left) or Roche NC total antibody (Ab) levels (right) on viral neutralization test (NT) titers is mediated by Technozym RBD antibody concentrations. *…P<0.05, **…P<0.01, ****…P<0.0001, R^2^ … Coefficient of determination, c… total effect, c’… direct effect of NC antibody levels on NT titer.

**Figure 2:**
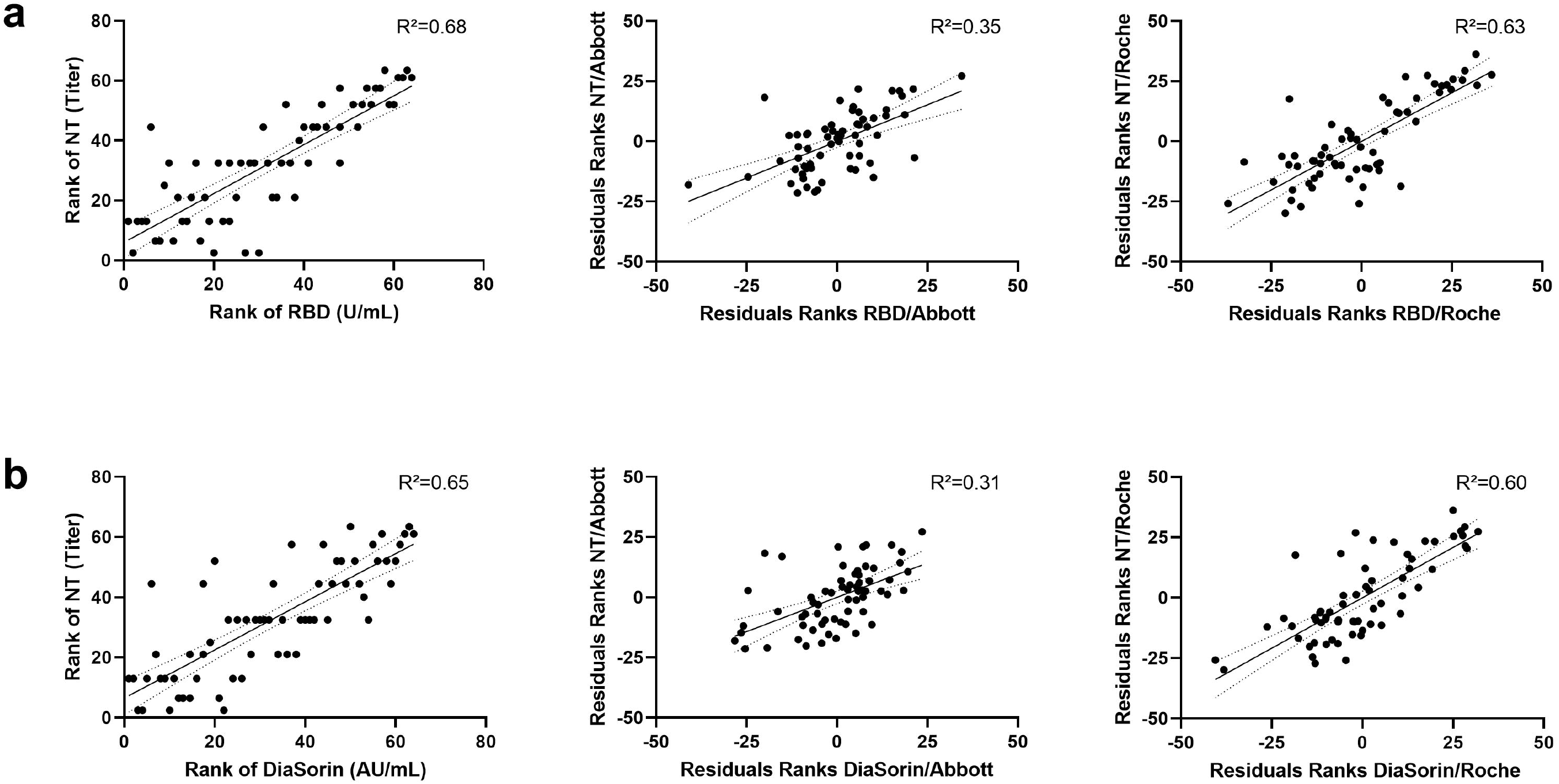
Rank correlations between titers of viral neutralization tests (NT) and a) RBD antibody concentrations quantified by the Technozym RBD ELISA or b) S1/S2 antibodies. The figures in the second and third column present rank correlations between NT and RBD with anti-nucleocapsid antibody concentrations kept constant in two different assays (Abbott, Roche). R^2^… Coefficient of determination.

## Conclusions

Our data suggest that the relationship between NC antibody levels and NT titers is only apparent and is mostly mediated by the concomitant presence of RBD or S1/S2 antibodies. Spike protein specific antibody concentrations and NT titers remain correlated after keeping NC antibody levels constant. In contrast, the correlation between NC antibody levels and NT titers nearly vanished for most of the assessed test systems after keeping RBD or S1/S2 antibody levels constant. This was especially true for Roche NC-assay measured total NC antibody levels, whereas the IgG-based assays appeared to correlate better with NT titers. The remaining weak correlation between the Abbott test results and the NT titers, while keeping RBD-or S1/S2 IgG antibody concentrations constant, suggests that a third mediator variable may be present.

To prevent falsely implied causal relationships between SARS-CoV-2 specific NC antibodies and neutralizing activity, all correlation analyses of non-spike-associated antibody assays and neutralization assays should include a partial correlation analysis to exclude a possible mediator effect of spike-associated antibodies.

## Supporting information

Graphical Abstract

## Data Availability

Data can be requested by interested researchers from the corresponding author.

## Acknowledgements

This study was conducted in cooperation with the MedUni Wien Biobank facility. We thank all sample donors for their contribution, as well as Miss Manuela Repl and Miss Marika Gerdov for perfect technical assistance.

## References

1. Gavor E, Choong YK, Er SY, Sivaraman H, Sivaraman J. Structural Basis of SARS-CoV-2 and SARS-CoV Antibody Interactions. Trends Immunol. 2020;41(11):1006–22.

2. Bal A, Pozzetto B, Trabaud MA, Escuret V, Rabilloud M, Langlois-Jacques C, et al. Evaluation of high-throughput SARS-CoV-2 serological assays in a longitudinal cohort of patients with mild COVID-19: clinical sensitivity, specificity and association with virus neutralization test. Clin Chem. 2021.

3. Brochot E, Demey B, Touzé A, Belouzard S, Dubuisson J, Schmit J-L, et al. Anti-spike, Anti-nucleocapsid and Neutralizing Antibodies in SARS-CoV-2 Inpatients and Asymptomatic Individuals. Frontiers in Microbiology. 2020;11(2468).

4. Tang MS, Case JB, Franks CE, Chen RE, Anderson NW, Henderson JP, et al. Association between SARS-CoV-2 neutralizing antibodies and commercial serological assays. Clin Chem. 2020.

5. McAndrews KM, Dowlatshahi DP, Dai J, Becker LM, Hensel J, Snowden LM, et al. Heterogeneous antibodies against SARS-CoV-2 spike receptor binding domain and nucleocapsid with implications for COVID-19 immunity. JCI Insight. 2020;5(18).

6. Laferl H, Kelani H, Seitz T, Holzer B, Zimpernik I, Steinrigl A, et al. An approach to lifting self-isolation for health care workers with prolonged shedding of SARS-CoV-2 RNA. Infection. 2020.

7. Reed LJ, Muench H. A SIMPLE METHOD OF ESTIMATING FIFTY PER CENT ENDPOINTS12. American Journal of Epidemiology. 1938;27(3):493–7.

8. Perkmann T, Perkmann-Nagele N, Breyer MK, Breyer-Kohansal R, Burghuber OC, Hartl S, et al. Side-by-Side Comparison of Three Fully Automated SARS-CoV-2 Antibody Assays with a Focus on Specificity. Clin Chem. 2020;66(11):1405–13.

9. Perkmann T, Perkmann-Nagele N, Oszvar-Kozma M, Koller T, Breyer M-K, Breyer- Kohansal R, et al. Increasing both specificity and sensitivity of SARS-CoV-2 antibody tests by using an adaptive orthogonal testing approach. medRxiv. 2020:2020.11.05.20226449.

10. Haslacher H, Gerner M, Hofer P, Jurkowitsch A, Hainfellner J, Kain R, et al. Usage Data and Scientific Impact of the Prospectively Established Fluid Bioresources at the Hospital- Based MedUni Wien Biobank. Biopreserv Biobank. 2018;16(6):477–82.

11. Hayes AF. Introduction to Mediation, Moderation, and Conditional Process Analysis. 2^nd^Ed. ed. New York: Guilford Press; 2018.

