## Supplementary figures and images for "Spike protein antibodies mediate the apparent correlation between SARS-CoV-2 nucleocapsid antibodies and neutralization test results"

### Graphical Abstract

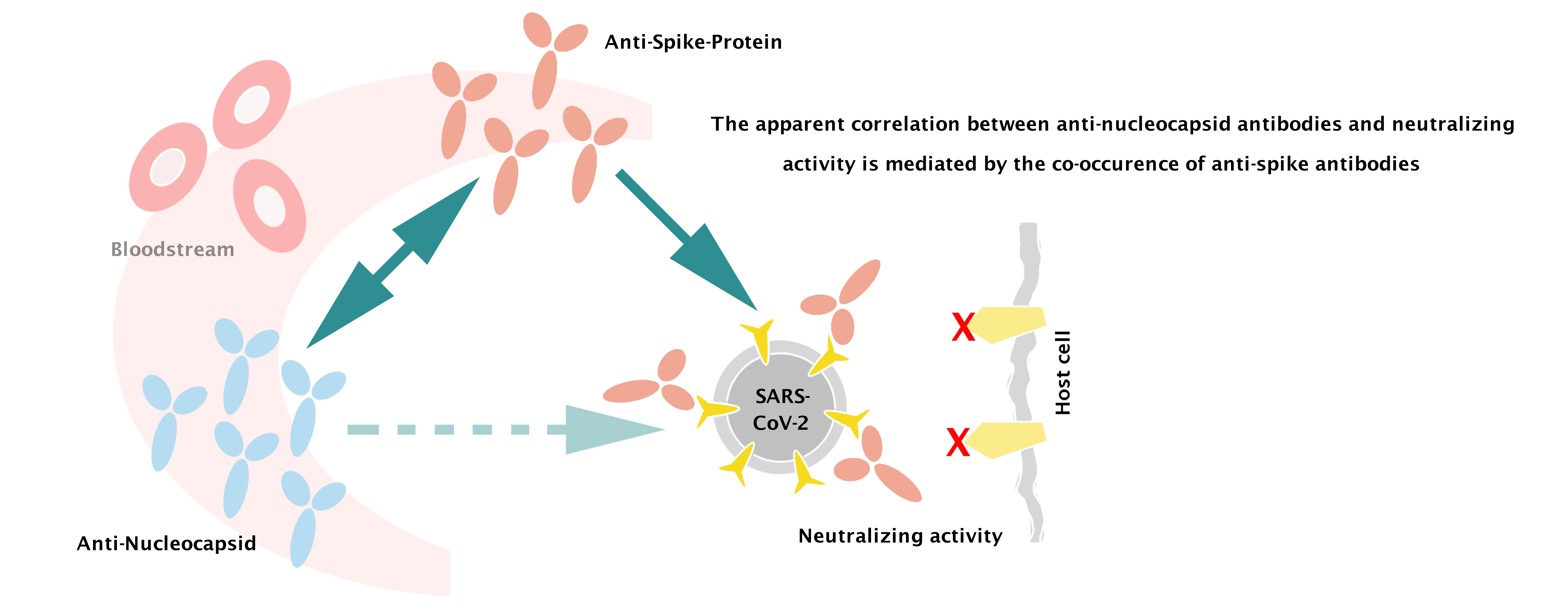
